# Cognitive impairment and functional change in COVID-19 patients undergoing inpatient rehabilitation

**DOI:** 10.1101/2021.03.15.21253637

**Authors:** Ruchi Patel, Irene Savrides, Christine Cahalan, Gargi Doulatani, Michael W. O’Dell, Joan Toglia, Abhishek Jaywant

## Abstract

Cognitive impairment is increasingly recognized as a sequela of COVID-19. It is unknown how cognition changes and relates to functional gain during inpatient rehabilitation. We administered the Montreal Cognitive Assessment (MoCA) at admission to 77 patients undergoing inpatient rehabilitation for COVID-19 in a large U.S. academic medical center. 45 patients were administered the MoCA at discharge. Functional gain was assessed by change in the Quality Indicator for Self-Care (QI-SC). In the full sample, 80.5% of patients exhibited cognitive impairment on admission, which was associated with prior delirium. Among 45 patients with retest data, there were significant improvements in MoCA and QI-SC. QI-SC score gain was higher in patients who made clinically meaningful changes on the MoCA, an association that persisted after accounting for age and delirium history. Cognitive impairment is frequent among COVID-19 patients, but improves over time and is associated with functional gain during inpatient rehabilitation.

## Introduction

Cognitive impairment is increasingly recognized as a sequela of mild and severe COVID-19 infection(1–4). However, it is still unclear how cognitive impairment after COVID-19 evolves and relates to functional outcomes. Cognitive may improve in COVID-19 patients undergoing rehabilitation(5,6), but existing studies have not evaluated the association between cognitive change and functional gain, particularly in inpatient rehabilitation when early interventions can be implemented. Cognition is associated with functional gain in rehabilitation in other illnesses(7–9). Determining whether such an association exists in the inpatient rehabilitation phase after COVID-19 could provide additional targets for early treatment to optimize outcome.

The primary goal of the present study was to evaluate change in cognition and its relation to functional gain among COVID-19 patients undergoing inpatient rehabilitation. A secondary goal was to add to a growing body of literature that describes the frequency, severity, and predictors of cognitive impairment after COVID-19.

## Methods

### Participants

94 patients with COVID-19 admitted to an inpatient rehabilitation unit in a United States medical center from March-August 2020. Patients were medically stable and able to tolerate 3 hours/day of occupational, physical, and speech-language therapy.

Seventeen patients were not administered our cognitive measure because (1) they were admitted prior to knowledge of COVID-19-related cognitive deficits and the implementation of our screening protocol; (2) clinicians deemed it could not be reliably administered due to language barriers; or (3) clinicians deemed patients were cognitively intact.

Data were extracted from retrospective chart reviews under approval of the Weill Cornell Medicine Institutional Review Board. 19/77 patients underwent a separate neuropsychological evaluation during their admission, the results of which have been published^5^; the results described here are novel.

### Outcome Measures

We extracted included age, gender (male or female), prior level of function (independent or not), intubation history (yes or no) and length (in days), delirium history (yes or no), and inpatient rehabilitation and acute hospitalization lengths of stay (Table 1).

**Table 1.** Demographic and clinical characteristics (N[%] or Mean[SD]).

|  | **N=77**<br>**(Full Sample)** | **N= 45**<br>**(Patients with**<br>**discharge**<br>**MoCA)** | **N=32**<br>**(Patients without**<br>**a discharge**<br>**MoCA)** |
| --- | --- | --- | --- |
| Gender |  |  |  |
| Male | 49 (63.6%) | 30 (66.7%) | 19 (59.4%) |
| Female | 28 (36.4%) | 15 (33.3%) | 13 (40.6%) |
| Age | 61.03 (15.67) | 61.98 (14.11) | 59.69 (17.77) |
| Independent Prior to Hospitalization | 74 (96.1%) | 44 (97.8%) | 30 (93.8%) |
| Acute hospitalization length of stay | 37.03 (31.8) | 39.4 (34.11) | 33.69 (28.4) |
| Inpatient rehabilitation unit length of stay | 13.43 (6.47) | 14.44 (6.06) | 12 (6.84) |
| Documented delirium | 44 (57.1%) | 28 (62.2%) | 16 (50%) |
| Intubated | 59 (76.6%) | 35 (77.8%) | 24 (75%) |
| Intubation length | 13.31 (10.53) | 13.75 (10.03) | 12.67 (11.37) |
| Documented hypoxia | 69 (89.6%) | 39 (86.7%) | 30 (93.8%) |
| History of neurological diagnosis | 2 (2.6%) | 1 (2.2%) | 1 (3.1%) |
| Initial MoCA scores | 20.29 (5.47) | 17.96 (4.71) | 23.47 (5.06) |
| Discharge MoCA scores |  | 22.02 (4.23) |  |
| Initial QI-SC score | 25.75 (6.21) | 24.96 (4.94) | 26.88 (7.59) |
| Discharge QI-SC score | 36.57 (8.37) | 37.11 (6.97) | 35.81 (10.09) |

The MoCA (10) was administered within 72 hours of admission to inpatient rehabilitation. Scores on the MoCA range from 0-30 and ranges indicate ≥26=normal, 18-25=mild impairment, 11-17=moderate impairment, and ≤10=severe impairment. The minimal clinical important difference (MCID) of 3 points was used(11). Three patients received the Blind MoCA, which was converted to 30-point scale per established guidelines. Discharge MoCA was administered to 45/77 patients. 32 patients did not receive a discharge MoCA as they either scored normal on the initial assessment, were unavailable, and/or declined.

The Quality Indicator for Self-Care (QI-SC) (12) is a standard functional assessment in the United States. Items scored on a scale of 1 to 6 include eating, grooming, toileting, bathing, upper body dressing, lower body dressing, and donning/doffing footwear. Total score ranges from 7 (total assistance) to 42 (independent). Occupational therapists assigned admission and discharge QI-SC scores to all patients.

### Statistical Analyses

We used descriptive statistics to describe the frequency of cognitive impairment. We used independent and paired-samples t-tests with effect sizes (Cohen’s d), and Spearman rank-order correlations, to evaluate associations between clinical characteristics and admission MoCA, change in MoCA and QI-SC from admission to discharge, and QI-SC change in patients who met or did not meet the MoCA MCID. To adjust for age and to account for the effect of multiple variables, we used multivariable linear regression models.

## Results

Table 1 provides demographic and clinical data. Patients with discharge MoCA scores (n=45) did not differ from those who were not administered the MoCA at discharge (n=32) in age, gender, prior neurologic history, prior delirium, prior intubation, or prior hypoxia (all p’s>.05).

### Cognitive Impairment at Admission to Rehabilitation

62/77 patients (80.5%) demonstrated cognitive deficits on the MoCA at admission: 39/77 (51%) had mild deficits, 20/77 (26%) had moderate deficits, and 3/77 (4%) had severe deficits.

### Correlates of Cognitive Impairment at Admission to Rehabilitation

Admission MoCA scores were lower for patients who previously had delirium (Mean Difference=3.08, 95% CI: .58-5.58, *t*=2.45, *p*=.02, *d*=.58). Admission MoCA scores were not correlated with age (r =-.15, *p*=.19), length of intubation (r = -.04, *p*=.72), or acute hospitalization length of stay (r =-.08 *p*=.50). In a multivariable linear regression predicting admission MoCA from age, history of delirium, length of intubation, and length of acute care hospitalization, the overall model was significant (*R*^*2*^=.13, *F*=2.53, *p*=.049). Among predictors, only history of delirium had a significant association with admission MoCA (β=-3.67, *t*=2.35, *p*=.02).

### Change in Cognition and Association with Functional Gain

The 45 patients with admission and discharge MoCA scores improved on the MoCA (Mean Difference=4.02, 95% CI: 2.92-5.13, *t*=7.35, *p*<.001, *d*=1.10) and QI-SC (Mean Difference=12.16, 95% CI: 10.3-14.01, *t*=13.19, *p*<.001, *d*=1.97) (Figure 1). 32/45 patients (71%) met the MoCA MCID. There was a greater increase in QI-SC scores in patients who met the MoCA MCID than those who did not (Mean Difference=4.41, 95% CI: 0.49-8.34, *t*=2.27, *p*=.03, *d*=0.75; Figure 2). To evaluate the robustness of this association, we conducted a multivariable linear regression with the outcome of QI-SC score change and predictors of MoCA MCID (met or did not meet/declined), age, and history of delirium. While the overall model was not statistically significant (*R*^*2*^=.13, *F*=2.03, *p*=.125), among individual predictors, improvement by the MoCA MCID continued to have a significant association with QI-SC score change (β=4.74, *t*=2.31, *p*=.03) after adjusting for age and history of delirium. Despite overall improvement, at discharge, 35/45 (78%) continued to exhibit cognitive impairment on the MoCA.

**Figure 1.**
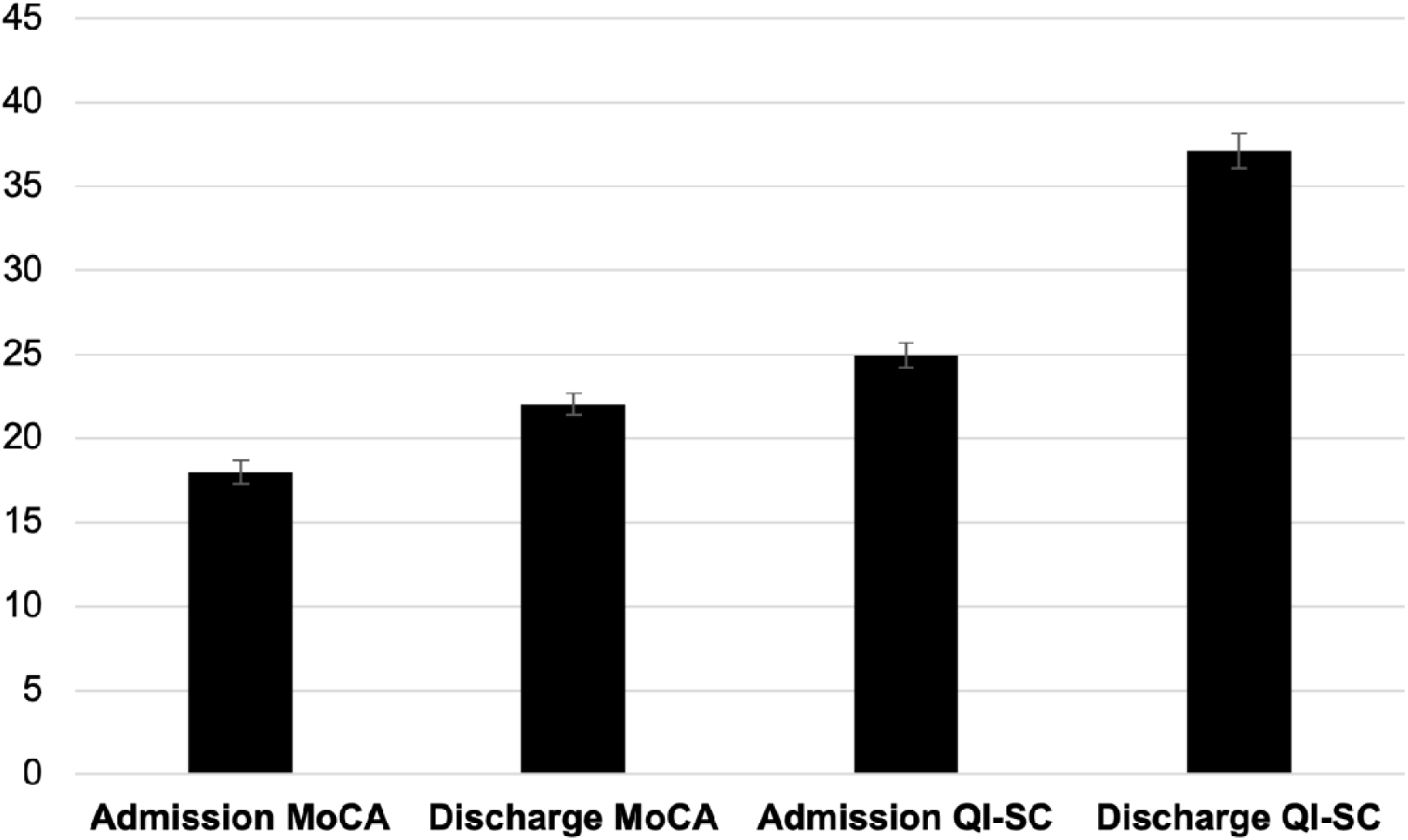
Mean scores on the MoCA and QI-SC at admission and discharge in N=45 patients with full data. Error bars represent SEM.

**Figure 2.**
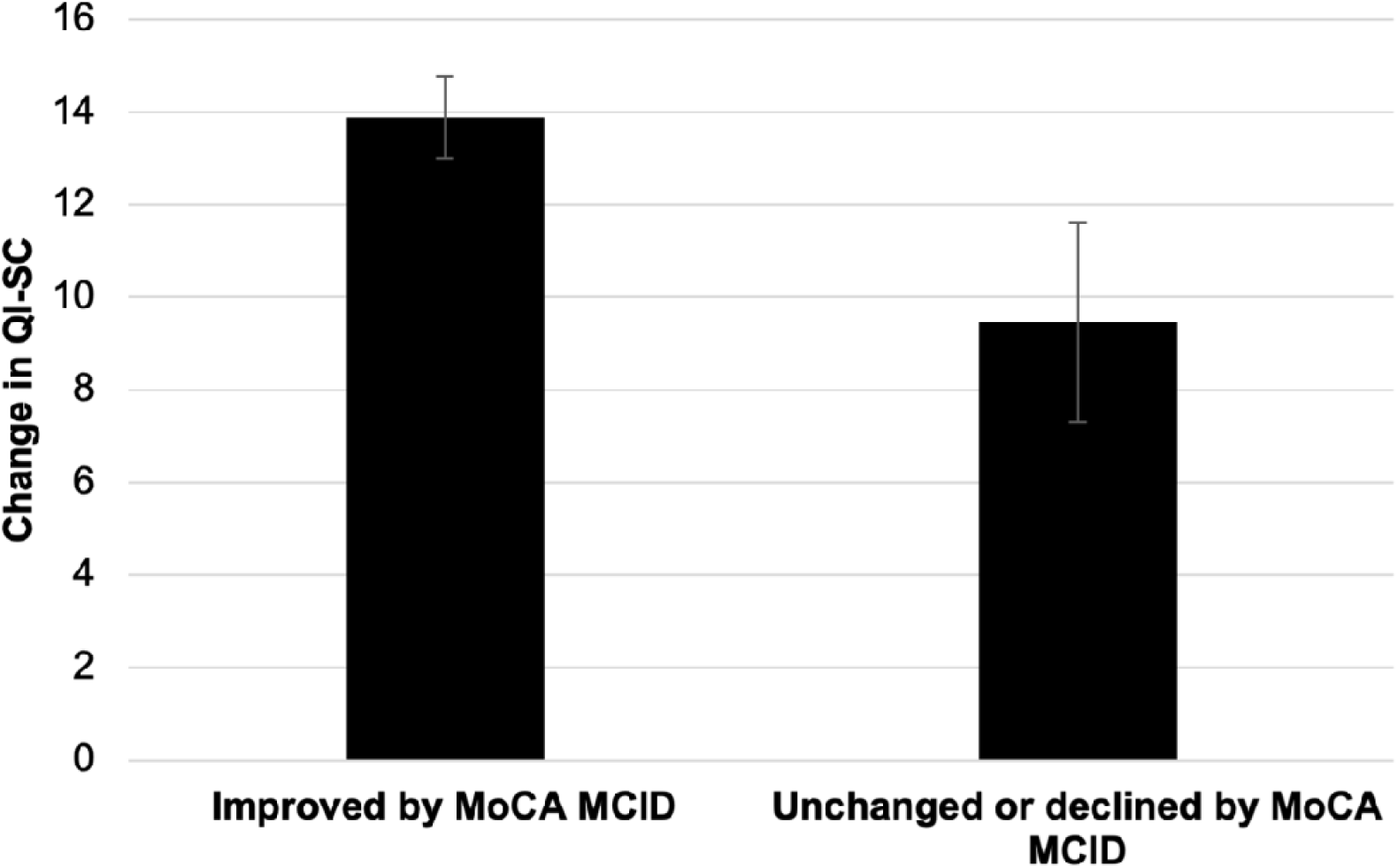
Change in functional self-care ability by clinically meaningful cognitive improvement. Error bars represent SEM.

## Discussion

Our main findings are (1) cognitive impairment is common in COVID-19 patients undergoing inpatient rehabilitation and associated with a history of delirium; (2) cognition and function (self-care) improves significantly during inpatient rehabilitation; and (3) clinically meaningful gains in cognition are associated with improved functional gains.

COVID-19 patients exhibited significant improvement in cognition with a large effect size. Cognitive improvement over time may reflect natural recovery, improvement in residual effects of prior delirium, and/or rehabilitation intervention effects. Our results indicate that cognition evolves after COVID-19 and suggests that it may be beneficial to implement cognitive interventions to facilitate functional recovery(5). Despite recovery, a majority of our sample continued to demonstrate cognitive impairment at discharge from inpatient rehabilitation, highlighting the importance of continued management of cognition post-discharge(6).

COVID-19 patients also experienced a significant improvement in their functional self-care skills. Notably, significantly greater functional improvement was seen in patients who exhibited a clinically meaningful improvement in cognition. Although we cannot make causal conclusions, these findings suggest that addressing cognitive impairment after COVID-19 may improve functional outcomes.

Consistent with prior research, we replicate findings of a high frequency of cognitive impairment, predominantly mild impairment, in recovering COVID-19 patients. Also consistent with the literature on the negative consequences of delirium after COVID-19(13–15), delirium during acute hospitalization was associated with greater cognitive impairment at admission to rehabilitation. This emphasizes the potential importance of early detection and management of delirium after COVID-19.

Among limitations of this study, discharge cognitive data were not available for 32/77 patients, which may have biased findings. However, there were no demographic or clinical differences between the 45 patients with and 32 patients without discharge cognitive data. Missing data highlight challenges of administering objective cognitive assessments in an acute inpatient setting, particularly during a time in which the cognitive sequelae of COVID-19 were relatively unknown and hospital policies and procedures were rapidly evolving at the initial surge of the pandemic. Prospective longitudinal research is necessary to estimate the true prevalence of cognitive dysfunction, its evolution post-hospitalization, and to ascertain etiological mechanisms. Our sample comprised a relatively sick group of patients from the first wave of COVID-19; findings may not apply to milder disease presentations or from subsequent waves of illness.

## Data Availability

Data for this manuscript are not available.

## Acknowledgements

We thank the staff of the IRU for assisting in cognitive and functional assessments, and for caring for, our COVID-19 patients.

